# Work Requirements and Health: Projected Mortality Impacts of Medicaid Coverage Loss

**DOI:** 10.1101/2025.02.28.25322887

**Authors:** Abhishek Pandey, Yang Ye, Burton H. Singer, Alison P. Galvani

## Abstract

Recent legislative proposals to impose nationwide work requirements for Medicaid eligibility could jeopardize health insurance coverage for over 36 million low-income Americans. To assess the potential mortality impact, we estimated annual excess deaths under a comprehensive set of scenarios reflecting varying policy implementations. These scenarios accounted for differences in age groups (19–55 vs. 19–64), eligibility based on Medicaid expansion status, and varying levels of automatic exemptions and reporting compliance. In the most optimistic scenarios—where work requirements are applied only to Medicaid expansion enrollees—we project 4,936 and 10,851 excess deaths annually among individuals aged 19–55 and 19–64, respectively. Under the most regressive scenario, where work requirements are imposed on all non-disabled Medicaid enrollees aged 19–64, the estimated annual excess deaths rise sharply to 53,144. These projections underscore the grave public health consequences of implementing Medicaid work requirements and highlight the need for careful evidence-based policymaking.

---

Recent proposals by congressional leaders to impose work requirements on Medicaid eligibility could jeopardize access to health care for 36.2 million Americans.^1^ Medicaid, a joint federal and state program, provides health insurance to low-income individuals and families. The Affordable Care Act (ACA), through its Medicaid expansion provision enabled states to broaden eligibility, significantly decreasing uninsured rates in participating states.

Many of those who will be affected already face significant socioeconomic challenges and a disproportionate disease burden, raising concerns that coverage losses would exacerbate health issues for those most in need. These requirements, often framed as initiatives to promote self-sufficiency and reduce dependence on public assistance, mandate that beneficiaries engage in a specified number of work hours, job training, or other qualifying activities to maintain their Medicaid coverage. Some proposals, including the Limit, Grow, Save Act, target enrollees aged 19 to 54 years^2^, while others extend the range to 18 to 65 years.^1^ In Arkansas and New Hampshire, where such requirements were briefly implemented, they applied only to individuals enrolled through the Medicaid expansion program. However, recent proposals do not clearly define such distinctions—suggesting that the entire Medicaid population could be affected. Most Medicaid enrollees already work,^3^ yet these regulations impose administrative burdens and unnecessary overhead expenses for taxpayers. Furthermore, the retractions will interrupt chronic care for many, potentially leading to more severe and costly health outcomes. Onerous reporting processes and administrative hurdles risk disenrollment even for those meeting work requirements or qualifying for exemptions. These challenges are compounded for enrollees with limited digital literacy, unstable work schedules, or difficulty navigating bureaucratic systems.^4^

Attempts at work requirements in Arkansas and New Hampshire have proven to be counterproductive. In Arkansas, 25% of Medicaid enrollees lost coverage within the first six months. Of those, 89% remained unenrolled after a year, often citing confusion over reporting rules and difficulties accessing the online portal.^5^ Similarly, New Hampshire suspended work requirements when it became apparent that thousands of individuals were at risk of losing coverage due to reporting bureaucracy.^6^ These examples highlight the potential for rapid, widespread disenrollment with lasting health repercussions.

We applied our previously established methodology^7^ to project the potential increase in annual deaths resulting from proposed policy changes, stratifying our analysis by age (19–55 and 19–64) and eligibility status (Medicaid expansion vs. all non-disabled). For scenarios only considering expansion enrollees, automatic exemption and reporting rates (rate of compliance or seeking exemption) were informed by data from Arkansas and New Hampshire.^8^ When modeling scenarios that included all Medicaid enrollees, we accounted for the high potential exemption rate among traditional enrollees (due to parental/caregiving responsibilities) by considering reporting requirements for everyone, thereby eliminating the effect of automatic exemptions. We also evaluated worst-case scenarios, where both exemption and reporting rates were assumed to be zero.

We first estimated the number of enrollees who would lose coverage due to the implementation of work requirements under each scenario considered. These figures were then used to update insurance coverage within the affected age group. Next, we applied the elevated mortality risk^9^ associated with being uninsured to estimate the resulting number of excess deaths (Supplementary Appendix). To reflect uncertainty in our projections, we incorporated empirical variability in both mortality hazard ratios and current insurance coverage, generating uncertainty intervals for our excess death estimates.

Our analysis projects a range of potential outcomes, with thousands of excess deaths estimated across 16 different policy scenarios (**Table 1**). Among expansion enrollees aged 19–55, assuming a 52% exemption rate, we project 4,936 excess annual deaths [95% UI: 1,014–10,128], corresponding to a 28% reporting rate based on the experience in Arkansas. Under a lower reporting rate of 18%, as observed in New Hampshire, excess deaths increase to 5,622 [95% UI: 1,155–11,534]. If the policy were applied to expansion enrollees aged 19–64, the corresponding excess deaths would be 10,851 [95% UI: 2,221–22,360] under a 28% reporting rate and 12,358 [95% UI: 2,530–25,466] under an 18% reporting rate.

**Table 1.** Excess annual deaths under scenarios imposing work requirements by age group and enrollment pathway.

| Non-disabled Medicaid enrollees subjected to work requirement |  |  |  |  |  |  | Excess annual deaths |  |  |
| --- | --- | --- | --- | --- | --- | --- | --- | --- | --- |
| Age-group | Group | Number | Automatic Exemption | Reporting rate | Percent losing insurance | People losing insurance | Median | 95% UI |  |
| 19–55 | Expansion | 16,136,512 | 52% | 18% | 39% | 6,289,992 | 5,622 | 1,155 | 11,534 |
|  |  |  |  | 28% | 34% | 5,522,919 | 4,936 | 1,014 | 10,128 |
|  |  |  | 0% | 18% | 82% | 13,231,940 | 11,826 | 2,429 | 24,264 |
|  |  |  |  | 28% | 72% | 11,618,289 | 10,384 | 2,133 | 21,305 |
|  |  |  |  | 0% | 100% | 16,136,512 | 14,422 | 2,962 | 29,590 |
|  | All | 28,805,648 | 0% | 18% | 82% | 23,620,631 | 21,110 | 4,336 | 43,314 |
|  |  |  |  | 28% | 72% | 20,740,067 | 18,536 | 3,807 | 38,032 |
|  |  |  |  | 0% | 100% | 28,805,648 | 25,744 | 5,288 | 52,822 |
| 19–64 | Expansion | 20,272,000 | 49% | 18% | 42% | 8,414,946 | 12,358 | 2,530 | 25,466 |
|  |  |  |  | 28% | 36% | 7,388,733 | 10,851 | 2,221 | 22,360 |
|  |  |  | 0% | 18% | 82% | 16,623,040 | 24,412 | 4,998 | 50,305 |
|  |  |  |  | 28% | 72% | 14,595,840 | 21,435 | 4,388 | 44,170 |
|  |  |  |  | 0% | 100% | 20,272,000 | 29,771 | 6,095 | 61,348 |
|  | All | 36,188,000 | 0% | 18% | 82% | 29,674,160 | 43,578 | 8,922 | 89,801 |
|  |  |  |  | 28% | 72% | 26,055,360 | 38,264 | 7,834 | 78,849 |
|  |  |  |  | 0% | 100% | 36,188,000 | 53,144 | 10,880 | 109,513 |

When extending the policy to all non-disabled Medicaid enrollees, the impacts become even more severe (**Table 1**). For the 19–55 group, moderate scenarios result in 18,536 [95% UI: 3,807–38,032] excess deaths, rising to 25,744 [95% UI: 5,288–52,822] in the worst-case scenario. In the broader 19–64 population, we project as many as 53,144 [95% UI: 10,880–109,513] annual excess deaths when all 36.2 million at-risk individuals lose coverage.

These findings underscore the insidious public health ramifications of Medicaid work requirements. Policymakers must cautiously weigh the objective of workforce participation against the paramount importance of preserving health insurance continuity, as well as the substantial logistical costs and implementation complexities. Legislators should rigorously consider these mortality impacts when evaluating such counterproductive policies.

## Data Availability

All publicly available data used for the study along with computational code written in Python Jupyter's notebook is publicly available at https://github.com/abhiganit/wr4medicaid

## Notes

### Competing Interest Statement

The authors have declared no competing interest.

### Funding Statement

APG acknowledges support from Burnett Endowment. The funding source had no influence on methodology, analysis, or interpretation of the results.

### Summary of Updates

In our revised manuscript, we have extended our analysis to evaluate 16 policy considerations compared to 2 before.

