## Supplementary Appendix for "Work Requirements and Health: Projected Mortality Impacts of Medicaid Coverage Loss"

Recent proposals to implement work requirements for Medicaid recipients have emerged at both federal and state levels.[1] These policies mandate that certain non-disabled, working-age Medicaid beneficiaries participate in employment or other sanctioned activities to retain coverage.

Using our previously developed method,[2] we estimated the number of annual excess deaths among Americans who may lose coverage under such policies. Our analysis considered a comprehensive set of policy design variations:

1. Age groups affected (19–55 vs. 19–64)
2. Eligibility groups affected (expansion only vs. all non-disabled enrollees)
3. Varying automatic exemption and reporting compliance rates

For scenarios involving only the expansion population, we applied an automatic exemption rate of 52% for 19–55[3] and 49% for 19–64.[4] We also examined scenarios without automatic exemptions, in which work requirements apply to all non-disabled Medicaid beneficiaries. Following the experiences from Arkansas and New Hampshire, we consider scenarios where 28% and 18%, respectively, of those subject to requirements either report compliance or applied for exemptions[3]. We also consider a worst-case scenario of 0% reporting compliance to generate an upper bound.

We denote the population aged 19–55 or 19–64 as P and the current number of annual deaths in this population as $D_c$ . We inform P using national population projection datasets from United States Census Bureau[5] and calculate $D_c$ by applying the latest death rate estimates.[6]

If the probability of death for insured individuals in population P is μ, then

$$D_c = \mu i_c P + \lambda \mu (1 - i_c) P,$$

where $i_c$ is the current insured proportion within this age group and λ is the hazard ratio of death among uninsured individuals compared to insured individuals. We used data from the United

States Census Bureau to inform the current insured proportion ($i_c$). The data provided point estimates along with margins of error corresponding to 90% confidence interval.[7] By sampling age-specific population and age-specific insured counts from their corresponding normal distribution, we generated 1,000 samples for the age-specific insurance coverage proportion. Given that being uninsured in the US is associated with a 40% (95% CI: 6%,84%) higher risk of death compared to insured individuals of the same age,[8] we constructed a normal distribution informed by the estimated mean and variance to sample 1,000 inputs for λ.

As Medicaid work requirements are enforced, the insurance coverage among individuals in population P would change. We calculate the updated insurance coverage as:

$$i_n = i_c - \frac{n}{P},$$

where n is the number of Medicaid beneficiaries in population P who would become uninsured, informed by the scenario considered.

Since the proportion of insured individuals in the impacted population changes from $i_c$ to $i_n$ due to policy implementation, the expected number of annual deaths becomes:

$$D_n = \mu i_n P + \lambda \mu (1 - i_n) P,$$

or equivalently:

$$D_n = \frac{i_n + \lambda (1 - i_n)}{i_c + \lambda (1 - i_c)} D_c$$

The additional annual deaths are then calculated as $D_n - D_c$.

All publicly available data used for the study, along with the computational code written in Python for data analysis, is publicly available.[9]
